# Diagnostic Accuracy of Artificial Intelligence in Classifying HER2 Status in Breast Cancer Immunohistochemistry Slides and Implications for HER2-Low Cases: A Systematic Review and Meta-Analysis

**DOI:** 10.1101/2024.11.04.24316688

**Authors:** Daniel Arruda Navarro Albuquerque, Matheus Trotta Vianna, Luana Alencar Fernandes Sampaio, Andrei Vasiliu, Eduardo Henrique Cunha Neves Filho

## Abstract

Breast cancer with overexpression of the Human Epidermal Growth Factor Receptor 2 (HER2) accounts for 15-20% of cases and is associated with poor outcomes. Although trastuzumab-deruxtecan (T-DXd) has traditionally demonstrated survival benefits in metastatic HER2-positive patients, the DESTINY-Breast04 trial expanded its effectiveness to those with immunohistochemistry (IHC) scores of 1+, and 2+ with negative in situ hybridisation, a subset of patients that has since been termed “HER2-low”. Accurate differentiation of HER2 scores has now become crucial. However, visual IHC scoring is labour-intensive and prone to high interobserver variability. AI has emerged as a promising tool in diagnostic medicine, particularly within histopathology. This study assesses AI’s ability to identify patients eligible for T-DXd and its performance in accurately classifying HER2 scores. Electronic searches were conducted in MEDLINE, EMBASE, Scopus, and Web of Science up to May 2024. Eligibility criteria were limited to studies evaluating the performance of AI compared to pathologists in classifying HER2 utilising IHC slides. Metaanalysis was performed using the bivariate random-effects model to estimate pooled sensitivity, specificity, concordance, and area under the curve (AUC). To explore sources of heterogeneity, subgroup analysis and meta-regression were performed. Risk of bias was assessed using QUADAS-AI tool. We analysed 25 contingency tables across thirteen included publications, showing excellent AI accuracy in predicting T-DXd eligibility, with a pooled sensitivity of 0.97 [95%CI 0.96-0.98], specificity of 0.82 [95%CI 0.73-0.88], and AUC of 0.98 [95%CI 0.96-0.99]. In the individual scores analysis, AI performed better particularly in scores 2+ and 3+. Substantial heterogeneity was observed, and meta-regression revealed better performance with deep learning and patch-based analysis, while performance declined in externally validated and those utilising commercially available algorithms. Our findings indicate that AI holds promising potential in accurately identifying HER2-low patients and excels in distinguishing 2+ and 3+ scores. Upcoming validation studies should focus on enhancing AI’s precision in the 0-1+ range and improving the reporting of clinical and pre-analytical data to standardise samples characteristics, ensuring models are more comparable to each other. This review highlights that deep learning advancements are driving automation, requiring pathologists to adapt and integrate this technology into their workflow.

## 1. Introduction

Breast cancer is the leading cause of cancer among women worldwide and is expected to result in approximately 1 million deaths per year by 2040 [1]. Between 15% and 20% of cases exhibit overexpression of the Human Epidermal Growth Factor Receptor 2 (HER2) [2], which is associated to poor clinical outcomes. The HER2 is a transmembrane tyrosine kinase receptor often found in breast cancer cells, and its activation promotes cell cycle progression and proliferation [3]. HER2 expression is routinely assessed semi-quantitatively by pathologists using immunohistochemistry (IHC)-stained slides, where HER2 is categorised based on the intensity and completeness of membrane staining into negative (scores 0 and 1+), equivocal (score 2+), or positive (score 3+). Equivocal cases undergo further testing with *in situ* hybridization (ISH), and are subsequently classified as either positive or negative depending on HER2 amplification status [2].

Historically, the use of antibody-drug conjugates, such as trastuzumab-deruxtecan (T-DXd), has been recommended for HER2-positive patients with metastatic disease, corresponding to those with IHC scores of 3+, or 2+ with gene amplification confirmed by ISH. Patients with scores of 0, 1+, or 2+ with negative ISH were typically eligible for clinician’s choice chemotherapy [4]. However, findings from the DESTINY-Breast04 (DB-04) [5] trial demonstrated that T-DXd significantly improved both progression-free and overall survival in individuals with low expression of HER2, with the latter particularly remarkable in the context of metastatic disease. The eligibility criteria for low HER2 expression in that study required an IHC score of 1+, or 2+ with negative ISH, in patients with metastatic breast cancer. This benefit in survival, observed in a subset of patients categorised as “HER2-low”, a designation introduced by the DB-04 trial, has prompted the American Society of Clinical Oncology/College of American Pathologists (ASCO/CAP) to emphasise the importance of distinguishing between scores 0 and 1+ [6], while maintaining the 2018 [2] four-class classification system.

Approximately 60% of HER2-negative metastatic breast cancers express low levels of HER2 [5]. Accurate differentiation between scores 1+/2+/3+ and 0 has become critical, as it influences clinical decision-making and affects the number of patients eligible for T-DXd.

Nevertheless, the standard IHC visual scoring of HER2 is time-consuming, labour-intensive, and subject to high interobserver variability [7, 8]. Digital pathology has emerged as a promising tool in medical diagnostics, aiding in the diagnosis and grading of cancers such as breast, lung, liver, skin, and pancreatic cancers [9]. To address the subjective nature of IHC visual scoring, artificial intelligence (AI) and deep learning (DL) have become powerful options for analysing histological specimens, particularly for HER2 scoring [10].

Briefly, the workflow of an AI-based HER2 scoring model involves pre-processing digitalised whole slide images (WSI) by fragmenting the images into smaller patches and selecting a region of interest (ROI) likely to contain relevant neoplastic tissue. These patches are then split into a learning and testing subsets to train and evaluate the algorithm’s performance, respectively. An internal validation process is subsequently performed to assess the model’s accuracy using WSIs from the same training dataset. To assess the algorithm’s reproducibility, some models undergo external validation, where performance is tested with an independent external dataset. The final outcome is the overall HER2 score, which is derived from the scoring aggregation of each individual patch [10].

To the best of the author’s knowledge, only one metaanalysis, conducted by Wu et al. [11], has been published on the use of AI for HER2 scoring in breast cancer. However, this review included a limited number of studies, leading to performance metrics with a wide margin of uncertainty. Additionally, sources of heterogeneity were not explored, given the small sample size.

In line with the challenges associated with HER2 scoring and the advancements in AI, we conducted an updated and clinically focused meta-analysis aimed at: (1) evaluating the performance of AI compared to pathologists’ visual scoring in accurately identifying patients eligible for T-DXd based on HER2 IHC score; and (2) assessing the accuracy of AI in classifying each individual HER2 score.

## 2. Materials and Methods

### 2.1. Protocol registration and study design

This systematic review adhered to the Preferred Reporting Items for Systematic Reviews and Meta-Analysis for Diagnostic Test Accuracy (PRISMA-DTA) guidelines (Table S1) and was registered in the International Prospective Register of Systematic Reviews (PROSPERO) under the identification number CRD42024540664.

### 2.2. Eligibility criteria

Inclusion in this meta-analysis was limited to studies that fulfilled all of the following criteria: (1) studies that utilised primary or metastatic breast cancer tissues in the form of digitalised WSIs, from female patients at any age, irrespective of tumour stage, histological subtype and hormone receptor status, employing conventional 3,3’-diaminobenzidine (DAB) staining; (2) studies evaluating the performance of AI algorithms as an index test; (3) studies comparing these algorithms to pathologists’ visual scoring as the reference standard; and (4) studies that reported AI performance metrics. Only original research articles published in English in peer-reviewed journals were included. No limitations were imposed on the publication date.

Studies were excluded if they lacked sufficient data to build a 2 × 2 contingency table of true positives (TP), false positives (FP), true negatives (TN), and false negatives (FN) across each HER2 category (0, 1+, 2+, and 3+). Additionally, studies that combined 0/1+ scores were excluded, given the clinical significance of this distinction for the purposes of this review. Studies conducted using animal tissues, those employing haematoxylin and eosin (H&E) staining, multi-omics approaches, as well as reviews, letters, preprints, and conference abstracts, were excluded.

### 2.3. Literature search

A comprehensive literature search was conducted in MEDLINE, EMBASE, Scopus, and Web of Science from inception up to 3 May 2024 using the following terms: (“artificial intelligence” OR “machine learning” OR “deep learning” OR “convolutional neural networks”) AND “breast cancer” AND “HER2”. We also examined the reference lists of studies to identify relevant articles. Two independent authors screened titles, abstracts, and full texts for eligibility. Disagreements were resolved through consensus with a senior pathologist.

### 2.4. Data extraction

The following data were collected using a pre-designed spreadsheet: study characteristics (author, year, and country), participant details (tumour features, sample size, data unit, and dataset), index test (algorithm specifications, deep learning, transfer learning, autonomy, external validation, type of internal validation, and the use of commercially available (CA) algorithms), reference standard (manual scoring and classification system), and 2 × 2 contingency tables for TP, FP, FN, and TN for each 0, 1+, 2+, and 3+ score. Supplementary data were requested from corresponding authors when necessary. Performance metrics were calculated using three distinct data units: cases, WSIs, and patches, which varied across the included studies. A second independent reviewer cross-checked all the collected data. If studies provided multiple contingency tables representing various outcomes for a single AI algorithm, only the data reflecting the best performance were collected. In cases where studies presented results evaluating different types of automation (assisted *vs*. automated) or different data units for the same algorithm, these tables were treated as independent.

### 2.5. Outcomes

The primary outcome was the AI pooled sensitivity, specificity, area under the curve (AUC), and concordance in distinguishing HER2 scores of 1+/2+/3+ from 0. The cut-off for positivity was defined as a score of at least 1+, since the decision to initiate T-DXd treatment is based on this threshold. The secondary outcome was to estimate the same pooled performance metrics for each individual score of 1+, 2+, and 3+ compared to their counterparts.

Each study contributed with at least four contingency tables, corresponding to the performance of each individual score. Importantly, the term “positivity,” as used here, applies solely to defining the cut-off for the meta-analysis and is not related to the histological classification of HER2.

### 2.6. Statistical analysis

Meta-analysis was conducted using the bivariate randomeffects model to calculate the pooled sensitivity and specificity, as significant heterogeneity was expected. Summary receiver operating characteristics (SROC) curves with 95% confidence interval (CI) and 95% prediction region (PR) were employed for visual comparisons and AUC estimation. Heterogeneity was evaluated using the Higgins inconsistency index statistic (I^2^), with 50% defined as moderate and ≥75% defined as high. Pooled concordance was calculated using a random effects meta-analysis of proportions, following the DerSimonian and Laird method. Potential sources of heterogeneity were explored through subgroup analysis and meta-regression, utilising covariates that were most likely to contribute to heterogeneity. A “leave-one-out” sensitivity analysis was performed by sequentially excluding one study at a time. Publication bias was evaluated using Deek’s funnel plot asymmetry test. Statistical analysis was conducted using STATA (version 17.0, StataCorp LLC, Texas, USA) with midas, metandi and metaprop modules. To assess the threshold effect, the Spearman correlation coefficient was calculated using Meta-DiSc (version 1.4, Ramón y Cajal Hospital, Madrid, Spain). A *p* < 0.05 was considered significant for the meta-analysis, meta-regression, and threshold effect evaluation. For the assessment of publication bias, a significance cut-off of *p* < 0.10 was applied.

### 2.7. Quality assessment

The risk of bias and applicability concerns in the included articles were meticulously evaluated independently by two authors using the adapted Quality Assessment Tool for Artificial Intelligence-Centered Diagnostic Test Accuracy Studies (QUADAS-AI) [12] across four domains: patient selection, index test, reference standard, and flow and timing. The detailed questionnaire is provided in Table S2.

## 3. Results

### 3.1. Study selection and characteristics

Our search yielded a total of 1,581 records published from inception to April 2024, which were reduced to 751 after the removal of 830 duplicates. One relevant study [19] was found through article’s bibliography search. The remaining records were then screened for relevance based on their titles and abstracts. Subsequently, 72 records were deemed suitable for inclusion and were assessed in full text. Of these, 59 studies were excluded, with the reasons summarised in Figure 1. Publications that grouped 0/1+ scores together and those that reported accuracy metrics solely as percentages without providing raw data were excluded. Corresponding authors were contacted in both instances, but failed to provide the necessary data. Finally, a total of 13 studies were included in this review for metaanalysis.

**Figure 1:**
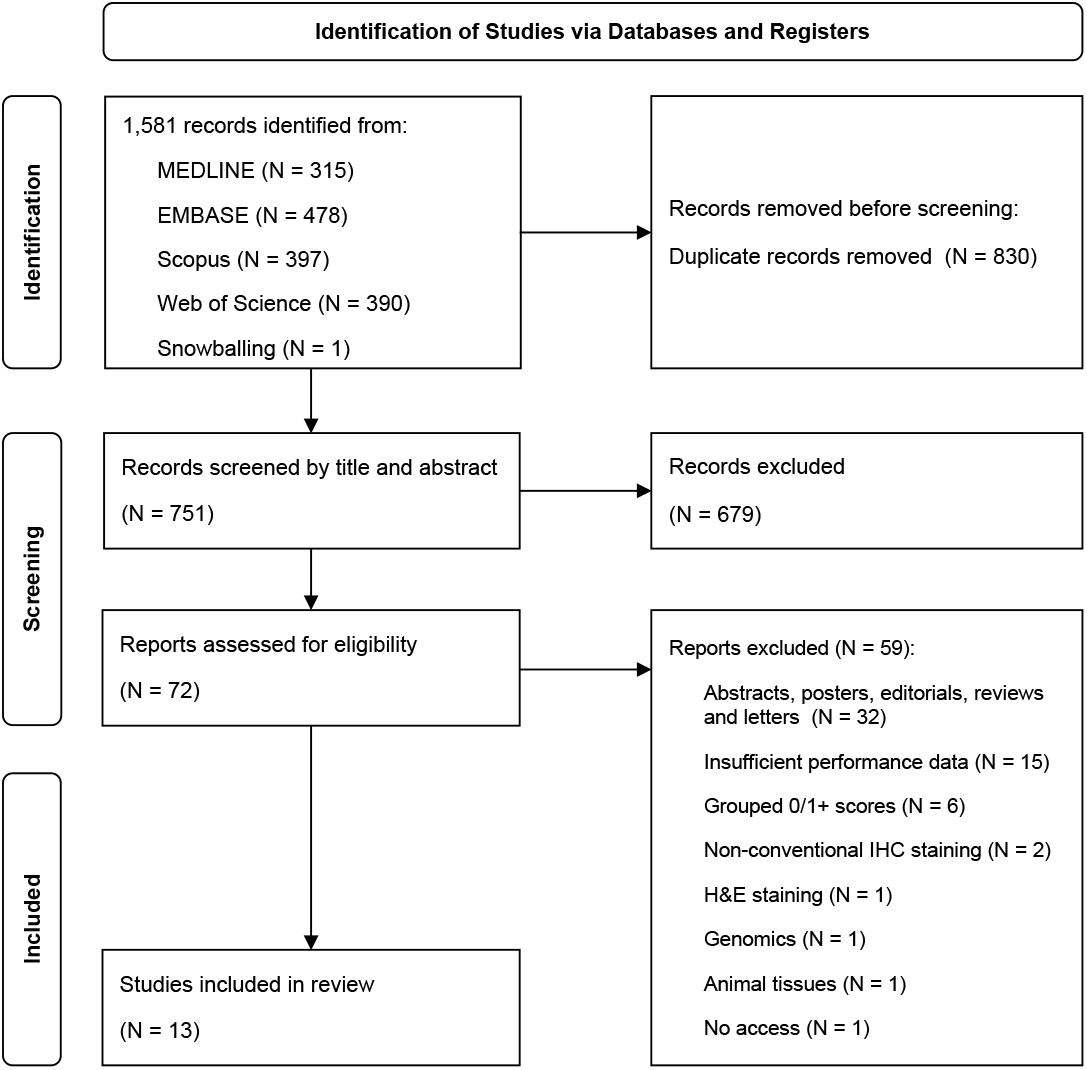
PRISMA flow diagram. H&E, haematoxylin and eosin; IHC, immunohistochemistry.

Table 1 presents a summary of the included studies published between 2018 and 2024, encompassing 1,285 cases, 168 WSIs, and 24,626 patches collected from 25 contingency tables. Only Palm et al. [20] and Sode et al. [23] specified the use of primary tumours, while the others did not mention the source. All authors reported invasiveness status, except for [14]. The type of tissue extraction (biopsy *vs*. resection) was reported in only five studies. The IHC assay was described in five studies, and [14] failed to give scanner details. The discrepancies in sample sizes observed in eight studies are attributable to the use of patches as the data unit, as multiple patches can be derived from a single WSI. Eight studies utilised, either wholly or in part, the HER2 Scoring Contest HER2SC [22] database, which may have resulted in some overlap of sample sizes. However, quantification of this was not possible, as authors did not specify which cases were used in their analyses. Ten studies employed DL, all of which utilised convolutional neural networks (CNN) algorithms. Only two studies exclusively utilised pathologist-assisted algorithms. In Pham et al. [25], pathologists manually selected a ROI before AI classification, while in Sode et al. [23], the final scoring required a pathologist’s review. Two studies [15, 20] tested the same algorithm for both automated and pathologist-assisted modes. One contingency table from [22] was excluded, as it used H&E-stained images as inputs for training the algorithm. Oliveira et al. [19] reported performance metrics using patches obtained from IHC-stained WSIs as an input for further prediction of HER2 status on H&Estained WSIs, and as such, it was included in this meta-analysis.

**Table 1:** Characteristics of the 13 included studies based on 25 contingency tables. ASCO, American Society of Clinical Oncology; CAP, College of American Pathologists; CNN, convolutional neural networks; HER2, Human Epidermal Growth Factor Receptor 2; HER2SC, HER2 Scoring Contest; NR, not reported; WSI, whole slide images.

| First author and year | Participants | Sample size | Data unit | Index test | Reference standard | Deep learning | Transfer learning | Autonomy | Internal validation | External validation | Commercially available | Dataset |
| --- | --- | --- | --- | --- | --- | --- | --- | --- | --- | --- | --- | --- |
| Bórquez et al. (2023) [13] | 86 cases of invasive breast carcinoma | 41<br>2898 | WSIs<br>Patches | Bayesian deep learning with Monte Carlo Dropout (CNN) | ASCO/CAP 2013 | Yes | No | Fully automated | 5-fold cross-validation | No | No | HER2 Scoring Contest (HER2SC) |
| Fan et al. (2024) [14] | 104 cases of breast tumour with overexpressed HER2 | 4000 | Patches | CNN | Average score of three pathologists (NR) | Yes | No | Fully automated | Random split-sample validation | No | No | Nanjing Medical University |
| Jung et al. (2024) [15] | 201 breast cancer patients | 201 | Cases | CNN: DeepLabv3+<br>ResNet-34<br>DeepLabv3<br>ResNet-101 | ASCO/CAP 2023 | Yes | Yes | Fully automated<br>Assisted | Random split-sample validation | Yes | No | Cureline (Brisbane, CA, US)<br>Superbiochips (Seoul, Republic of Korea)<br>Kyung Hee University Hospital (Seoul, Republic of Korea) |
| Kabir et al. (2024) [16] | 86 cases of invasive breast carcinoma | 77<br>5626 | WSIs<br>Patches | CNN: DenseNet201<br>GoogLeNet<br>MobileNetV2<br>Vision Transformer | ASCO/CAP 2018 | Yes | Yes | Fully automated | 5-fold cross-validation | No | No | HER2SC |
| Mirimoghaddam et al. (2024) [17] | 86 cases of invasive breast carcinoma<br>126 cases of invasive ductal breast carcinoma | 3200 | Patches | CNN: InceptionResNetV2<br>InceptionV3 | ASCO/CAP 2018 | Yes | Yes | Fully automated | 5-fold cross-validation | No | No | HER2SC<br>Ghaem Educational Research and Treatment Centre (Mashhad University) |
| Mukundan (2019) [18] | 86 cases of invasive breast carcinoma | 1206 | Patches | Multi-class logistic regression | ASCO/CAP 2018 | No | No | Fully automated | 5-fold cross-validation | No | No | HER2SC |
| Oliveira et al. (2020) [19] | 52 cases of invasive breast cancer | 2495 | Patches | CNN | ASCO/CAP 2018 | Yes | No | Fully automated | Random split-sample validation | No | No | HER2SC |
| Palm et al. (2023) [20] | Core needle biopsies of 67 primary invasive breast cancer | 67<br>201 | Cases<br>Patches | HER2 4B5® (Roche, Switzerland) | ASCO/CAP 2018 | No | No | Fully automated<br>Assisted | NR | Yes | Yes | Pathologie Institute Enge (Zurich, Switzerland) |
| Pedraza et al. (2024) [21] | 86 cases of invasive breast carcinoma<br>52 cases of invasive ductal carcinomas<br>15 cases of invasive and *in situ* ductal carcinomas | 5000 | Patches | CNN: ResNet-101<br>DenseNet-201 | ASCO/CAP 2018 | Yes | Yes | Fully automated | Random split-sample validation | Yes | No | HER2SC<br>Servicio de Salud de Castilla-La Mancha<br>Servicio Andaluz de Salud |
| Pham et al. (2023) [21] | 86 cases of invasive breast carcinoma | 50 | WSIs | CNN: U-Net<br>DenseNet121<br>ImageNet<br>ResNet18<br>ImageNet | ASCO/CAP 2018 | Yes | Yes | Assisted | 4-fold cross-validation | Yes | No | HER2SC<br>Erasmus Hospital |
| Qaiser et al. (2018) [22] | 86 cases of invasive breast carcinoma | 28 | Cases | MUCS-1 team: AlexNet (CNN)<br>VISILAB team: GoogLeNet (CNN) | ASCO/CAP 2018 | Yes | Yes | Fully automated | Random split-sample validation | No | No | "Men versus Machine" event of HER2SC |
| Sode et al. (2023) [23] | 727 cases of invasive primary breast carcinoma | 761 | Cases | HER2-CONNECT® (Vistopharm, Denmark) | ASCO/CAP 2018 | No | No | Assisted | NR | Yes | Yes | Zealand University Hospital |
| Yao et al. (2022) [24] | 228 biopsies of invasive breast carcinoma of no special type | 228 | Cases | GrayMap + CNN | ASCO/CAP 2018 | Yes | No | Fully automated | 5-fold cross-validation | No | No | Peking University Cancer Hospital & Institute |

### 3.2. Pooled performance of AI and heterogeneity

To assess the clinical relevance of AI in determining eligibility for T-DXd, we evaluated its performance in distinguishing scores 1+/2+/3+ from score 0 with data obtained from 25 contingency tables. When the threshold of positivity was set at score 1+, 2+, or 3+, with score 0 considered negative, the meta-analysis showed a pooled sensitivity of 0.97 [95% CI 0.96 - 0.98], a pooled specificity of 0.82 [95% CI 0.73 - 0.88], and an AUC of 0.98 [95% CI 0.96 - 0.99] (Figure 2).

**Figure 2:**
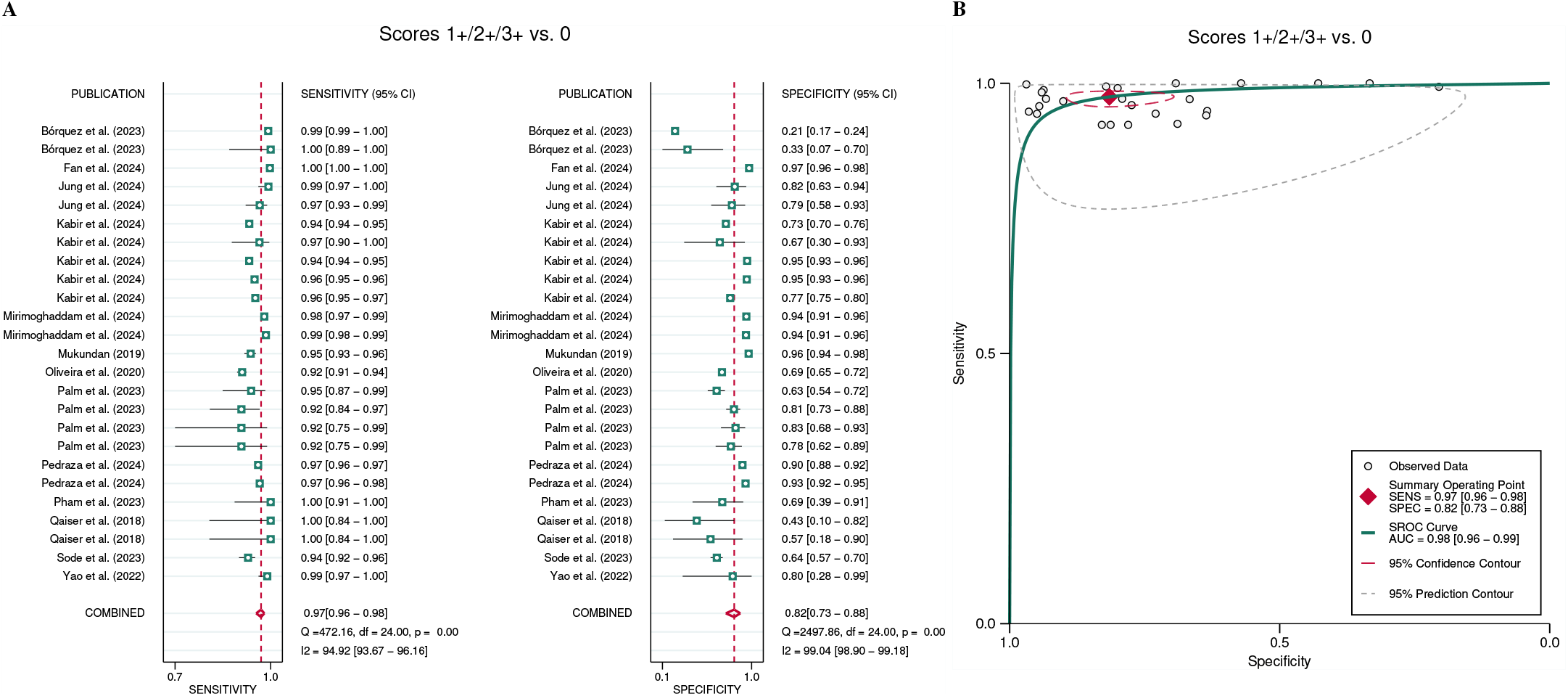
Pooled performance of AI in identifying HER2-low individuals from 25 contingency tables. **(A)** Forest plot of paired sensitivity and specificity for HER2 scores 1+/2+/3+ *vs*. 0. **(B)** Summary receiver operating characteristics curves for HER2 scores 1+/2+3+ *vs*. 0. AUC, area under the curve; SENS, sensitivity, SPEC, specificity

We also examined the performance of AI in distinguishing scores 1+, 2+, and 3+ individually from their counterparts. In this case, the test was considered positive if the respective score was identified by the models and negative if any other score was detected. Figures 3 and S1 present the corresponding SROC curves and forest plots, respectively. Notably, the performance improved with higher HER2 scores (2+ and 3+). The pooled sensitivity for score 1+ was 0.69 [95% CI 0.57 - 0.79], with a specificity of 0.94 [95% CI 0.90 - 0.96] and an AUC of 0.92 [95% CI 0.90 - 0.94]. For score 2+, AI achieved a pooled sensitivity of 0.89 [95% CI 0.84 - 0.93], a pooled specificity of 0.96 [95% CI 0.93 - 0.97], and an AUC of 0.98 [95% CI 0.96 - 0.99]. Finally, for score 3+, the AI demonstrated near-perfect performance, with a sensitivity of 0.97 [95% CI 0.96 - 0.99], specificity of 0.99 [95% CI 0.97 - 0.99], and an AUC of 1.00 [95% CI 0.99 - 1.00].

**Figure 3:**
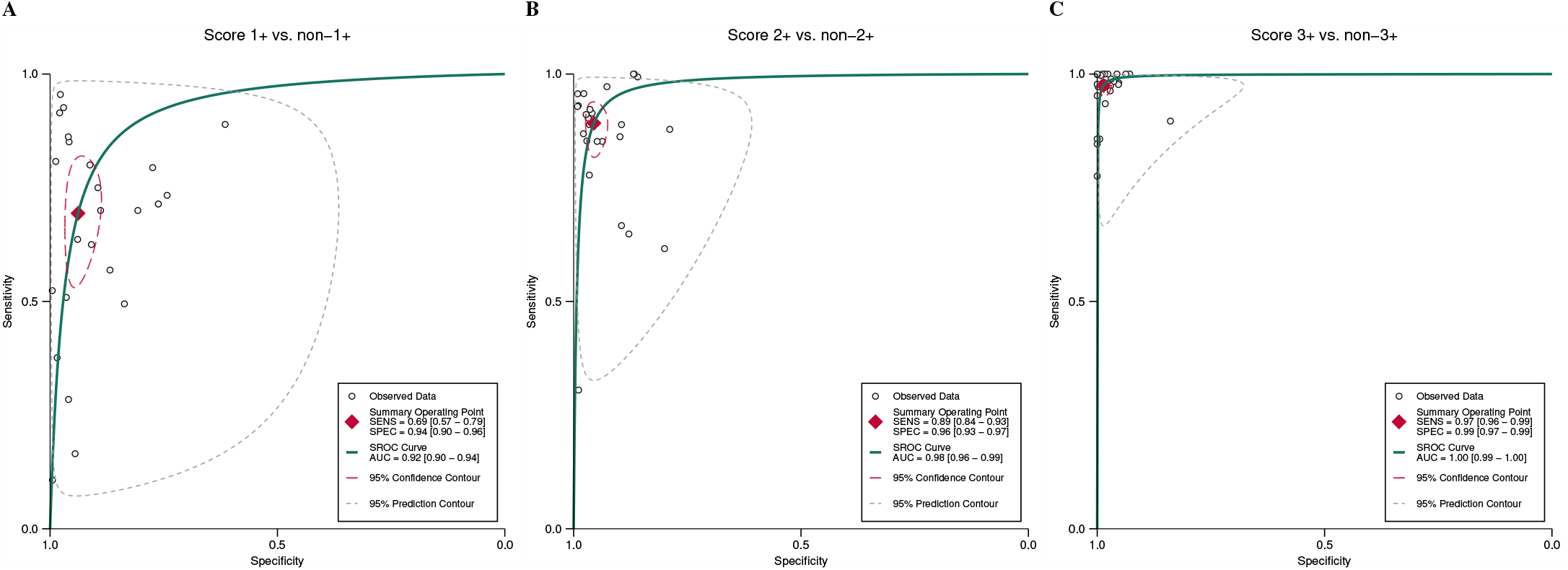
Summary receiver operating characteristics curves for individual HER2 scores. **(A)** score 1+ *vs*. non-1+, **(B)** score 2+ *vs*. non-2+, and **(C)** score 3+ *vs*. non-3. AUC, area under the curve; SENS, sensitivity, SPEC, specificity.

Our results indicate substantial heterogeneity, with I^2^ values ranging from 94% to 98% for both sensitivity and specificity (Figures 2 and S1). In all analyses, no significant threshold effect was identified (Table S3).

Figure 4 provides a heatmap with a visual representation of the agreement between AI and pathologists across all HER2 scores. The highest agreement was observed at score 3+, with a concordance of 97% [95% CI 96 - 98%], while AI performed less accurately at score 1+ (88% [95% CI 86 - 90%]).

**Figure 4:**
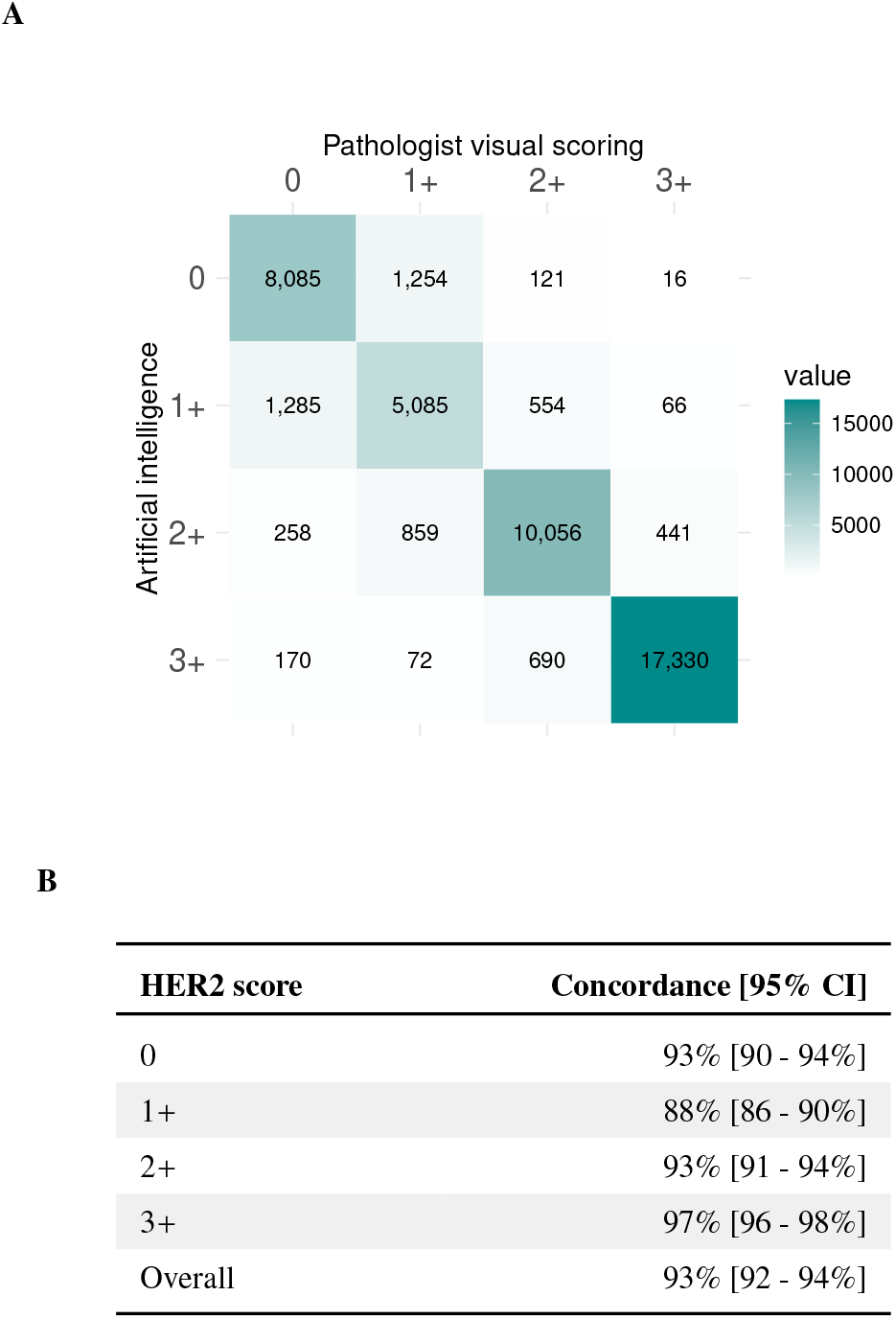
Concordance of AI *vs*. pathologists’ visual scoring. **(A)** Heatmap displaying the frequency of accurate and inaccurate AI predictions for each HER2 score. Cell values represent the cumulative sample sizes derived from 25 contingency tables corresponding to each prediction. **(B)** Concordance ratios of correct *vs*. incorrect predictions across different HER2 scores. CI, confidence interval; HER2, Human Epidermal Growth Factor Receptor 2.

### 3.3. Subgroup analysis and meta-regression

To investigate sources of heterogeneity in the 1+/2+/3+ *vs*. 0 analysis, we conducted meta-regression utilising potential covariates, including: (1) DL (present *vs*. not present); (2) transfer learning (present *vs*. not present); (3) CA algorithms (present *vs*. not present); (4) autonomy (assisted *vs*. automated); (5) type of internal validation (random split sample *vs. k*-fold crossvalidation); (6) external validation (present *vs*. not present); (7) sample size (≤761 *vs*. >761, where 761 is the median across studies); (8) data unit (WSIs/cases *vs*. patches); and (9) dataset (own *vs*. HER2SC). The performance estimates corresponding to each covariate and the meta-regression results is outlined in Table S4. Two studies [20, 23] failed to provide information about the type of internal validation and were not included in this subgroup analysis.

Considering sensitivity alone, we identified statistically significant differences across studies using DL, CA algorithms, and external validation. Studies incorporating DL achieved greater sensitivity (0.98 [95% CI 0.97 - 0.99] *vs*. 0.94 [95% CI 0.93 - 0.95]). In contrast, CA algorithms exhibited lower sensitivity (0.93 [95% CI 0.90 - 0.95]) relative to experimental-only algorithms (0.98 [95% CI 0.97 - 0.99]). Similarly, studies employing external validation have also performed worse (Sensitivity of 0.96 [95% CI 0.95 - 0.97] *vs*. 0.98 [95% CI 0.96 - 0.99]) (Table S4).

Regarding specificity, only the covariates sample size and data unit demonstrated significant differences. Analyses with sample sizes greater than 761 showed improved specificity (0.88 [95% CI 0.81 - 0.93]) compared to those with sample ≤ sizes 761 (0.70 [95% CI 0.55 - 0.82]). When patches were used as the data unit, specificity was significantly higher (0.87 [95% CI 0.79 - 0.92]) in contrast to 0.70 [95% CI 0.53 - 0.83] for WSIs/cases (Table S4). The covariates transfer learning, autonomy, type of internal validation, and dataset did not show a statistically significant impact on either sensitivity or specificity.

### 3.4. Publication bias and sensitivity analysis

To assess publication bias, we performed Deek’s funnel plot asymmetry test for all threshold analyses. The studies showed reasonable symmetry around the regression lines, with a nonsignificant effect, indicating a low likelihood of publication bias (Figure S2). Sensitivity analysis revealed no significant changes in performance estimates for the 1+/2+/3+ *vs*. 0 metaanalysis (Table S5).

### 3.5. Quality assessment

The QUADAS-AI assessment of the included studies is summarised in Figure S3, with a detailed description of the questionnaire outlined in Table S2. Briefly, 11 studies demonstrated a high risk of bias in the “subject selection” domain due to unclear reporting of eligibility criteria, such as the modality of tissue extraction (biopsy *vs*. resection), tumour invasiveness, and failure to specify whether the tumours were primary or metastatic. In the same domain, two studies [20, 23] that utilised CA algorithms did not provide a clear rationale neither a detailed breakdown of their training and validation sets. The “index test” domain exhibited high risk of bias in eight studies, as they lacked external validation. Furthermore, eight studies that evaluated AI performance using patches were deemed to have high applicability concerns regarding their index tests, as the use of WSIs/cases instead, would be more representative of real-world practice.

## 4. Discussion

In this meta-analysis, we found that AI demonstrated high performance in distinguishing HER2 scores of 1+/2+/3+ from 0, a critical cut-off with significant clinical importance. Considering the 1+, 2+ and 3+ scores individually, the AI models showed increasing sensitivity and specificity as scores rose, with particularly strong performance at higher scores. For score 3+, AI had almost perfect concordance with pathologist’s visual scoring. Meta-regression revealed that the use of DL, larger sample sizes, and patches positively influenced AI performance, while the use of externally validated and CA models showed less favourable outcomes.

The classification of scores 0 and 1+ is often challenging for pathologists, leading to significant inter-observer variability. A survey by Fernandez et al. [8], conducted across 1,452 laboratories over two years, found that pathologists agreed on the differentiation of scores 0 *vs*. 1+ in only 70% of cases or fewer. Given the findings of the DB-04 trial, our analysis primarily focused on the distinction between scores 1+/2+/3+ and 0, as patients scoring 1+, 2+, or 3+ are potentially eligible for T-DXd therapy, irrespective of ISH amplification status. Clinically, it is crucial to minimise the number of patients who might miss the opportunity to benefit from T-DXd and those who might be inappropriately treated. The pooled performance metrics found in this work, however, may not be readily intuitive. Put simply, for every 100 patients diagnosed with metastatic breast cancer, AI missed three eligible patients for T-DXd therapy, while eighteen patients would be erroneously treated (Figure 2), experiencing the drug’s adverse effects without a clear therapeutic benefit. This is especially relevant given the potential life-threatening side-effects associated with antibody-drug conjugates. For instance, in the T-DXd arm of the DB-04 trial, 45 patients (12.1%) experienced drug-related interstitial lung disease or pneumonitis, leading to the death of three patients (0.8%).

A similar meta-analysis by Wu et al. [11], which assessed AI-assisted HER2 status based on four studies differentiating scores 1+/2+/3+ from 0, found a pooled sensitivity of 0.93 [95% CI 0.92 - 0.94], a pooled specificity of 0.80 [95% CI 0.56 - 0.92], and an AUC of 0.94 [95% CI 0.92 - 0.96]^1^. Despite the broad 95% CI observed in that review, we found a significantly higher sensitivity (Figure 2). In contrast, our findings for specificity were similar, and AUCs were borderline comparable (Figure 2(b)). The less availability of studies in their analysis likely explains the discrepancy in sensitivity. While they applied similar inclusion criteria and a broader search strategy, yielding four times more studies than this review, five of our included studies were published after their search period concluded. Interestingly, these studies were released after the 2023 ASCO/CAP updated recommendations on HER2-low [6] were made public, indicating a growing demand for more accurate AI models since then.

Meta-regression indicated that DL improved significantly the sensitivity in identifying patients eligible for T-DXd (Table S4). DL is an AI tool that offers higher accuracy and robustness compared to traditional machine learning (ML) models. It has been widely applied in diagnostic medicine, particularly in histopathology [26], due to its effectiveness in analysing visual data. In this review, all studies that employed DL used CNN, a type of DL model that automatically learns and extracts image features, requiring minimal human intervention. CNN can automatically identify features such as texture, colour intensity, and shape with little need of pathologists’ annotations or selection of ROIs [27]. In contrast, traditional ML models are heavily dependent on pathologists’ expertise and experience, usually requiring human inputs for maximum effectiveness. The higher sensitivity observed in the DL subgroup is likely explained by its ability to identify image features that may have been overlooked by the human eye. However, DL automation requires large datasets and high-performance computing infra-structure for optimal performance [28], which can be a limitation in practice, as laboratories may not be equipped with such resources.

We also found significantly better specificity in the subgroups with a sample size >761 and those that used patches as the primary unit of analysis. These results are likely interrelated, as both subgroups showed similar disparities in specificity (Table S4). Patches are small, localised segments of a larger WSI, designed to facilitate the analysis of WSIs with extremely high resolution [26], therefore studies that reported performance metrics using patches are expected to have larger sample sizes. In HER2 scoring, models typically analyse each patch separately and then aggregate the results into a patient-level overall score, taking into account spatial distribution and the relative weight of scores across individual patches. Although patches can be utilised at some point during AI training, the overall WSI score is the most applicable in practice. However, studies that reported performance outcomes using patches lacked an overall aggregated score. From a practical perspective, the higher specificity observed in the patches subgroup suggests that the real-world performance of these models may be overestimated, as the ability to reliably aggregate individual patch scores into a comprehensive patient-level score was not fully evaluated.

Studies that underwent external validation and utilised CA models showed poorer sensitivity. Since CA algorithms rely on pre-designed and pre-trained models, their testing datasets are inherently external to the training data, leading to similar outcomes across both subgroup analyses (Table S4). Indeed, to mitigate overfitting and ensure that models generalise beyond the training set, it is essential to use samples from an external dataset. This accounts for variations in WSIs across different settings, such as distinct staining protocols, scanner resolutions, and imaging artefacts. As a result, algorithms tested on the same training dataset were expected to perform significantly better, but not necessarily would be a viable option in practice. The individual analysis of scores 1+, 2+, and 3+ *vs*. their counterparts showed that sensitivity, specificity and concordance improved as scores increases (Figures 3, 4 and S1). This trend aligns with the similar meta-analysis by Wu et al. [11]. Unlike the analysis of T-DXd eligibility, where patient management and classification in HER2-low were clinician-driven, AI’s performance in classifying individual HER2 scores is more relevant in a histopathology context, as the classic four-class HER2 recommendations remain in effect [6]. The lower sensitivity in score 1+ may reflect AI’s limitations in detecting subtle immunohistochemical changes, particularly at the 10% threshold of faint membranes between scores 0 and 1+ as per ASCO/CAP [29, 2]. This may suggest that either the detection or counting of faint/barely perceptible membranes is downregulated, as most incorrect predictions were biased towards score 0 (Figure 4). Conversely, the high specificity for score 2+ would be beneficial for laboratory workflows, as for every 100 equivocal cases (2+), ISH would be erroneously performed in only four, sparing costs and avoiding delays in clinical decisionmaking.

The intra- and inter-observer concordance rates for IHC manual HER2 scoring vary significantly across the literature. Concordance studies differ in methodology, as pathologists’ performance can be assessed either through agreement with peers or with more advanced lab assays. Additionally, factors such as the number of raters, wash-out period, variations in IHC assays, and the accuracy of the reference standard test used as the ground truth introduce further heterogeneity to these studies. A multi-institutional cohort study conducted with 170 breast cancer biopsies across 15 institutions reported an overall four-class (0, 1+, 2+, and 3+) concordance among pathologists as low as 28.82% [95% CI 22.01 - 35.63%] [30]. In contrast, another study found a markedly better figure of 94.2% [95% CI 91.4 - 96.5%] [31]. Studies comparing pathologists’ performance in IHC *vs*. other techniques reported concordance rates of 93.3% [32], 91.7% [33], 82.9% [34] with RT-qPCR, and 82% [35] with ISH. Based on these metrics, the overall concordance rate of 93% [95% CI 92 - 94%] (Figure 4) identified in this review implies that AI may hold the potential to achieve performance comparable to pathologists’ visual scoring.

However, the findings of this work have limitations. The exclusion of studies that used grouped 0/1+ scores and those that did not report sufficient performance outcomes may have influenced our pooled metrics. This is likely because, in earlier studies, separating 0/1+ scores was not relevant for T-DXd eligibility at that time, and DL technology had not yet become as widespread and advanced as it currently is. The notably high heterogeneity found in our analyses suggests that AI may lack standardisation, due to inherent differences in models design as well as pre-analytical factors, such as IHC assay, WSI scanner resolution, tumour heterogeneity, and tissue artefacts. The design of a standardised testing protocol is likely to make the various AI models more comparable to each other. Moreover, the high risk of bias and applicability concerns identified in QUADAS-AI (Figure S3) represent an additional limitation. Some included studies failed to report important sample characteristics, such as tumour invasiveness, method of extraction (biopsy *vs*. resection), or whether tumours were metastatic or primary, we cannot extrapolate our findings to samples that differ in these aspects. For instance, it cannot be ignored that algorithms may perform differently if non-breast cells are present in metastatic samples or if slides varies in size depending on the method of extraction. It remains essential that upcoming validation studies in AI provide a more detailed description of the samples, taking pre-analytical clinical and factors into account.

In light of the current ASCO/CAP recommendations, the pooled performance findings in this meta-analysis apply to the HER2-low population, though ongoing research may prompt changes in the HER2 classification system. Despite the US Food and Drug Administration [36] and the European Medicines Agency [37] approvals of T-DXd for metastatic breast cancer following the DB-04 trial, it is widely agreed among ASCO/CAP and European Society of Medical Oncology (ESMO) that HER2-low does not represent a biologically distinct entity, but rather a heterogeneous group of tumours influenced by HER2 expression [6, 38]. We acknowledge that the 2023 ASCO/CAP update reaffirms the 2018 version of the HER2 scoring guideline, with no prognostic value attributed to HER2-low and no changes recommended in reporting HER2 categories. The term HER2-low was used as an eligibility criterion in the DB-04 trial and should guide clinical decisionmaking only. To date, HER2 classification remains as negative (0 and 1+), equivocal (2+), and positive (3+). This is primarily because the DB-04 trial did not include patients with score 0, and the possibility that some patients in the 0-1+ threshold spectrum might benefit from T-DXd should not be ignored. Addressing this uncertainty, the recently published phase 3 DESTINY-Breast06 [39] trial found a survival benefit in HER2-ultralow patients, i.e., a subset of patients within the IHC category 0 who express faint membrane staining in ≤10% of tumour cells. Accordingly, HER2-ultralow hormone receptor-positive patients who had undergone one or more lines of endocrine-based therapy had longer progression-free survival when treated with T-DXd compared to those receiving chemotherapy. These findings align with the interim outcomes of the ongoing phase 2 DAISY trial [40]. The grey zone within the 0-1+ threshold has several implications for this review’s outcomes. All AI algorithms included in this analysis used the conventional 4-class HER2 scoring system (0, 1+, 2+, 3+), and it is clear that this classification may need to be revised. Subsequent AI algorithms will need to be trained using inputs that account for staining intensity and the proportion of stained cells within the 0-1+ spectrum.

In conclusion, the results of this meta-analysis, encompassing 25 contingency tables across thirteen studies, suggest that AI achieved promising outcomes in accurately identifying HER2-low individuals eligible for T-DXd therapy. Our findings indicate that, to date, AI achieves substantial accuracy, particularly in distinguishing higher HER2 scores. Future validation studies using AI should focus on improving accuracy in the 0-1+ range and provide more detailed reporting of clinical and pre-analytical data. Lastly, this review highlights that the progressive development of DL is steering towards greater automation, and pathologists will need to adapt to effectively integrate this tool into their practice.

## Data Availability

All data produced are available online at MEDLINE, EMBASE, Scopus and Web of Science.

https://pubmed.ncbi.nlm.nih.gov

https://www.embase.com/landing?status=grey

https://www.scopus.com/home.uri

https://clarivate.com/academia-government/scientific-and-academic-research/research-discovery-and-referencing/web-of-science/

## Author contributions

D.A.N.A developed the study concept and design, conducted the electronic searches, performed data extraction, and wrote the first draft of this manuscript. M.T.V contributed to data analysis and interpretation, statistical review, and critical revision of the manuscript. A.V assisted with electronic searches, data extraction, and risk of bias assessment. L.A.F.S and E.H.C.N.F contributed to the conceptualisation, critical review, and supervision. All authors have approved the final version of this manuscript.

## Data availability

The datasets used and/or analysed in this study are provided in the supplementary materials. Additional datasets can be obtained from the corresponding author upon request.

## Funding

This study was carried out independently and has not been sponsored by any public or private organisation.

## Declaration of competing interest

The authors declare no conflicts of interest.

## Ethics approval and consent to participate

This research utilised publicly accessible data, thus ethical approval and informed consent were not applicable.

## Declaration of generative AI and AI-assisted technologies in the writing process

During the preparation of this work the authors used [Chat-GPT 4o version 1.2024.227] in order to improve readability and language. After using this tool, the authors reviewed and edited the content as needed and take full responsibility for the content of the publication.

## Supplementary Material Legends

## 1. PRISMA-DTA Checklist

**Table S1:** PRISMA-DTA checklist.

| Section/topic | # | PRISMA-DTA Checklist Item | Reported on page # |
| --- | --- | --- | --- |
| **TITLE / ABSTRACT** |  |  |  |
| Title | 1 | Identify the report as a systematic review (+/- meta-analysis) of diagnostic test accuracy (DTA) studies. | 1 |
| Abstract | 2 | Abstract: See PRISMA-DTA for abstracts. | 1 |
| **INTRODUCTION** |  |  |  |
| Rationale | 3 | Describe the rationale for the review in the context of what is already known. | 1, 2 |
| Clinical role of index test | D1 | State the scientific and clinical background, including the intended use and clinical role of the index test, and if applicable, the rationale for minimally acceptable test accuracy (or minimum difference in accuracy for comparative design). | 2 |
| Objectives | 4 | Provide an explicit statement of question(s) being addressed in terms of participants, index test(s), and target condition(s). | 2 |
| **METHODS** |  |  |  |
| Protocol and registration | 5 | Indicate if a review protocol exists, if and where it can be accessed (e.g., Web address), and, if available, provide registration information including registration number. | 2 |
| Eligibility criteria | 6 | Specify study characteristics (participants, setting, index test(s), reference standard(s), target condition(s), and study design) and report characteristics (e.g., years considered, language, publication status) used as criteria for eligibility, giving rationale. | 2 |
| Information sources | 7 | Describe all information sources (e.g., databases with dates of coverage, contact with study authors to identify additional studies) in the search and date last searched. | 2 |
| Search | 8 | Present full search strategies for all electronic databases and other sources searched, including any limits used, such that they could be repeated. | 2 |
| Study selection | 9 | State the process for selecting studies (i.e., screening, eligibility, included in systematic review, and, if applicable, included in the meta-analysis). | 2 |
| Data collection process | 10 | Describe method of data extraction from reports (e.g., piloted forms, independently, in duplicate) and any processes for obtaining and confirming data from investigators. | 2, 3 |
| Definitions for data extraction | 11 | Provide definitions used in data extraction and classifications of target condition(s), index test(s), reference standard(s) and other characteristics (e.g. study design, clinical setting). | 2, 3 |
| Risk of bias and applicability | 12 | Describe methods used for assessing risk of bias in individual studies and concerns regarding the applicability to the review question. | 3 |
| Diagnostic accuracy measures | 13 | State the principal diagnostic accuracy measure(s) reported (e.g. sensitivity, specificity) and state the unit of assessment (e.g. per-patient, per-lesion). | 3 |
| Synthesis of results | 14 | Describe methods of handling data, combining results of studies and describing variability between studies. This could include but is not limited to: a) handling of multiple definitions of target condition. b) handling of multiple thresholds of test positivity, c) handling multiple index test readers, d) handling of indeterminate test results, e) grouping and comparing tests, f) handling of different reference standards | 3 |
| Meta-analysis | D2 | Report the statistical methods used for meta-analyses, if performed. | 3 |
| Additional analyses | 16 | Describe methods of additional analyses (e.g., sensitivity or subgroup analyses, meta-regression), if done, indicating which were pre-specified. | 3 |
| **RESULTS** |  |  |  |
| Study selection | 17 | Provide numbers of studies screened, assessed for eligibility, included in the review (and included in meta-analysis, if applicable) with reasons for exclusions at each stage, ideally with a flow diagram. | 3 |
| Study characteristics | 18 | For each included study provide citations and present key characteristics including: a) participant characteristics (presentation, prior testing), b) clinical setting, c) study design, d) target condition definition, e) index test, f) reference standard, g) sample size, h) funding sources | 3, 4 |
| Risk of bias and applicability | 19 | Present evaluation of risk of bias and concerns regarding applicability for each study. | 6 |
| Results of individual studies | 20 | For each analysis in each study (e.g. unique combination of index test, reference standard, and positivity threshold) report 2x2 data (TP, FP, FN, TN) with estimates of diagnostic accuracy and confidence intervals, ideally with a forest or receiver operator characteristic (ROC) plot. | 5, 6 |
| Synthesis of results | 21 | Describe test accuracy, including variability; if meta-analysis was done, include results and confidence intervals. | 5, 6, 7 |
| Additional analysis | 23 | Give results of additional analyses, if done (e.g., sensitivity or subgroup analyses, meta-regression; analysis of index test: failure rates, proportion of inconclusive results, adverse events). | 6 |
| **DISCUSSION** |  |  |  |
| Summary of evidence | 24 | Summarize the main findings including the strength of evidence. | 6 |
| Limitations | 25 | Discuss limitations from included studies (e.g. risk of bias and concerns regarding applicability) and from the review process (e.g. incomplete retrieval of identified research). | 8 |
| Conclusions | 26 | Provide a general interpretation of the results in the context of other evidence. Discuss implications for future research and clinical practice (e.g. the intended use and clinical role of the index test). | 8 |
| **FUNDING** |  |  |  |
| Funding | 27 | For the systematic review, describe the sources of funding and other support and the role of the funders. | 9 |

### 2. Description of QUADAS-AI Risk of Bias and Applicability Concerns

**Table S2:** Description of QUADAS-AI questionnaire.

| Domain | Signalling question |
| --- | --- |
| **Subject selection** | **Risk of bias** |
|  | • Is the source, size, and quality of the input data clearly defined, along with patient eligibility criteria?<br>• Was the data obtained from non-open-source datasets?<br>• Is there a clear rationale and distribution provided for training, validation, and test sets?<br>• Was image pre-processing performed?<br>• Is the scanner model used for imaging acquisition specified? |
|  | **Applicability concerns** |
|  | • Are there concerns that the included patients and setting do not match the review question? |
| **Index test** | **Risk of bias** |
|  | • Was external validation conducted? |
|  | **Applicability concerns** |
|  | • Are there concerns that the index test, its conduct, or interpretation differ from the review question? |
| **Reference standard** | **Risk of bias** |
|  | • Is the reference standard likely to correctly identify the target condition? |
|  | **Applicability concerns** |
|  | • Are there concerns that the target condition as defined by the reference standard does not match the question? |
| **Flow and timing** | **Risk of bias** |
|  | • Was the time between conducting the index test and the reference standard appropriate? |

### 3. Forest Plots of Individual 1+, 2+, and 3+ HER2 Scores

**Figure S1:**
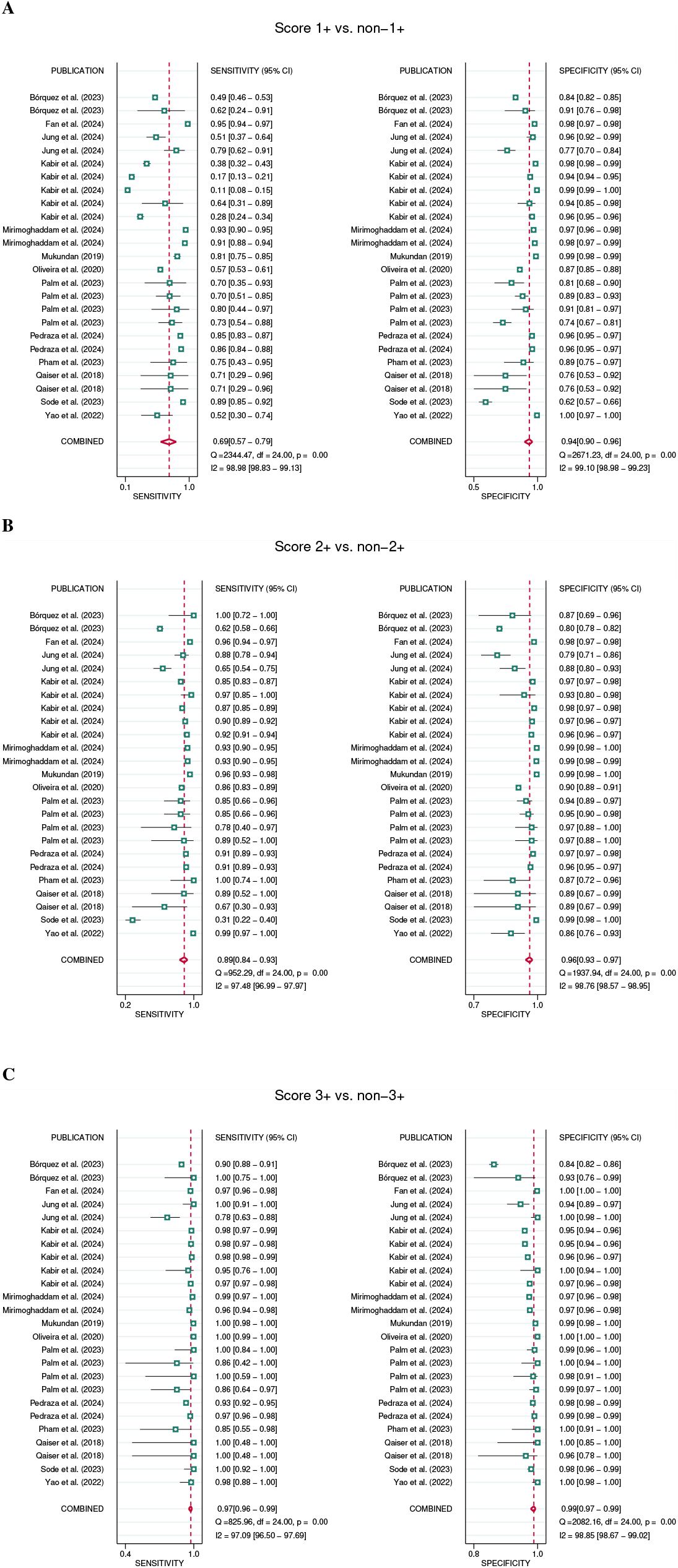
Forest plots of paired sensitivity and specificity for HER2 scores from 25 contingency tables. **(A)** score 1+ *vs*. non-1+, **(B)** score 2+ *vs*. non-2+, and **(C)** score 3+ *vs*. non-3+

### 4. Analysis of Threshold Effect

**Table S3:** Spearman correlation test. Coefficients were computed between sensitivity and specificity using logit transformations.

| HER2 cut-off | Spearman correlation coefficient | *p*-value |
| --- | --- | --- |
| 1+/2+/3+ vs. 0 | 0.050 | 0.811 |
| 1+ | 0.087 | 0.679 |
| 2+ | -0.068 | 0.746 |
| 3+ | 0.107 | 0.610 |

### 5. Subgroup Analysis and Meta-Regression of HER2 scores 1+/2+/3+ *vs*. 0

**Table S4:** Subgroup analysis and meta-regression of AI in distinguishing HER2 scores 1+/2+/3+ *vs*. 0. *p*-values were obtained from the likelihood ratio test comparing models with and without the covariates using mixed-effect logistic regression. “N” represents the number of contingency tables utilised in each subgroup. CI, confidence interval; HER2SC, HER2 Scoring Contest; WSI, whole slide images

| Covariate | Subgroup | Sensitivity [95% CI] | p-value | Specificity [95% CI] | p-value | N |
| --- | --- | --- | --- | --- | --- | --- |
| Deep learning | No | 0.94 [0.93 - 0.95] | $p < 0.001$ | 0.81 [0.67 - 0.90] | $p = 0.959$ | 6 |
|  | Yes | 0.98 [0.97 - 0.99] |  | 0.81 [0.71 - 0.89] |  | 19 |
| Commercially available algorithm | No | 0.98 [0.97 - 0.99] | $p = 0.001$ | 0.83 [0.72 - 0.90] | $p = 0.121$ | 20 |
|  | Yes | 0.93 [0.90 - 0.95] |  | 0.73 [0.64 - 0.81] |  | 5 |
| External validation | No | 0.98 [0.96 - 0.99] | $p = 0.006$ | 0.82 [0.67 - 0.91] | $p = 0.989$ | 15 |
|  | Yes | 0.96 [0.95 - 0.97] |  | 0.82 [0.74 - 0.88] |  | 10 |
| Sample size | ≤761 | 0.98 [0.95 - 0.99] | $p = 0.897$ | 0.70 [0.55 - 0.82] | $p = 0.018$ | 13 |
|  | >761 | 0.97 [0.96 - 0.98] |  | 0.88 [0.81 - 0.93] |  | 12 |
| Data unit | WSIs/cases | 0.98 [0.96 - 0.99] | $p = 0.370$ | 0.70 [0.53 - 0.83] | $p = 0.048$ | 11 |
|  | Patches | 0.97 [0.95 - 0.98] |  | 0.87 [0.79 - 0.92] |  | 14 |
| Transfer learning | No | 0.97 [0.95 - 0.99] | $p = 0.872$ | 0.76 [0.61 - 0.87] | $p = 0.229$ | 11 |
|  | Yes | 0.98 [0.96 - 0.99] |  | 0.85 [0.75 - 0.92] |  | 14 |
| Autonomy | Assisted | 0.97 [0.92 - 0.99] | $p = 0.607$ | 0.77 [0.53 - 0.91] | $p = 0.614$ | 5 |
|  | Automated | 0.98 [0.96 - 0.98] |  | 0.83 [0.73 - 0.89] |  | 20 |
| Type of internal validation | Random split sample | 0.98 [0.96 - 0.99] | $p = 0.492$ | 0.83 [0.66 - 0.93] | $p = 0.907$ | 8 |
|  | k-fold cross validation | 0.98 [0.96 - 0.99] |  | 0.82 [0.68 - 0.91] |  | 12 |
| Dataset | Own | 0.98 [0.96 - 0.99] | $p = 0.619$ | 0.82 [0.72 - 0.90] | $p = 0.833$ | 9 |
|  | HER2SC | 0.97 [0.96 - 0.98] |  | 0.81 [0.68 - 0.89] |  | 16 |

### 6. Analysis of Publication Bias

**Figure S2:**
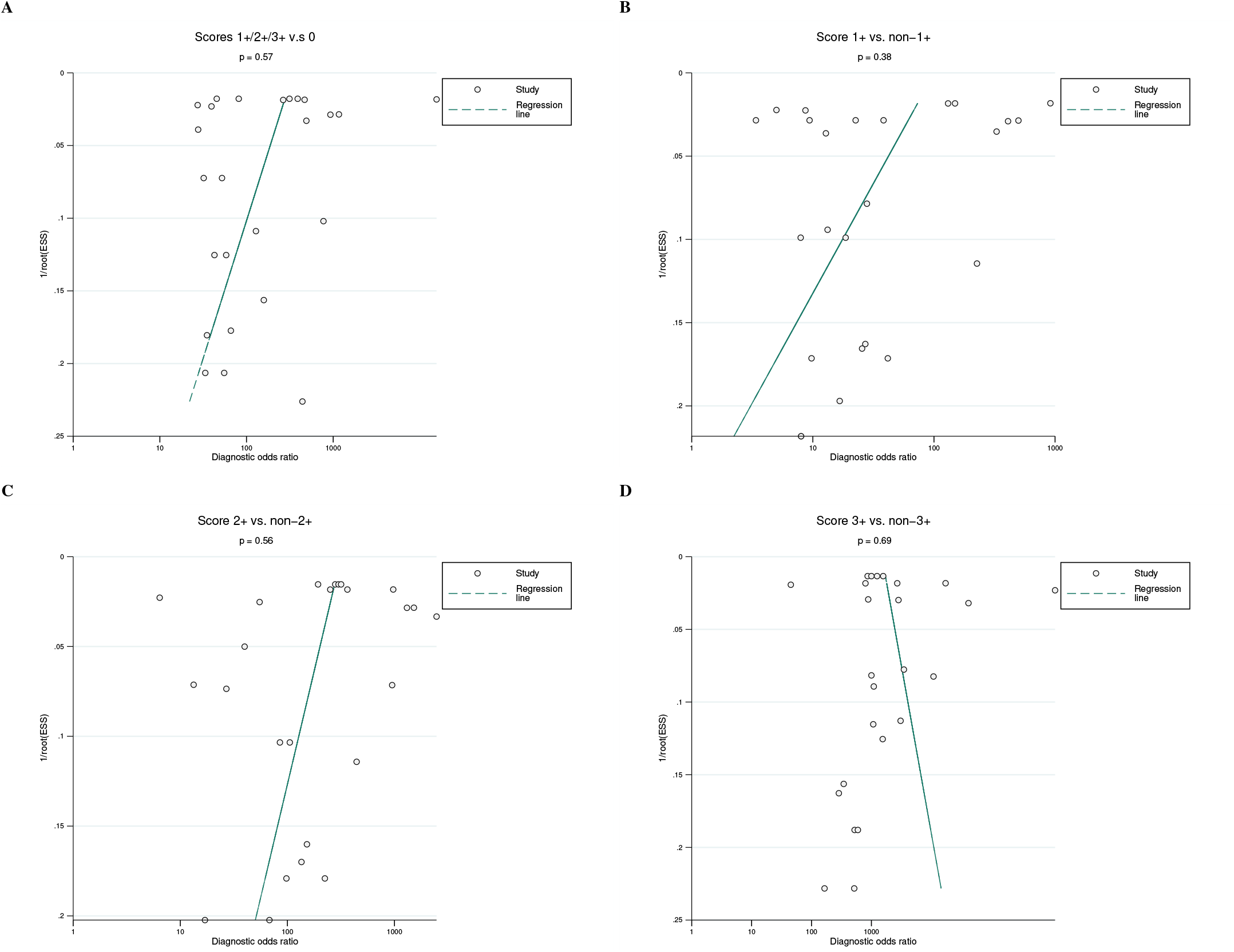
Deek’s funnel plot with superimposed regression line. The asymmetry test was performed using a regression of the diagnostic log odds ratio, weighted by the 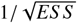. A *p* < 0.10 for the slope coefficient indicates significant asymmetry and high likelihood of publication bias. **(A)** scores 1+/2+/3+ *vs*. 0, **(B)** score 1+ *vs*. non-1+, **(C)** score 2+ *vs*. non-2+, and **(D)** score 3+ *vs*. non-3+. ESS, effective sample size.

### 7. Sensitivity Analysis

**Table S5:** Sensitivity analysis of the 1+/2+/3+ *vs*. 0 meta-analysis. Each row represents the performance when the corresponding study was excluded at a time from the overall meta-analysis. CI, confidence interval.

| Excluded study | Sensitivity [95% CI] | Specificity [95% CI] | AUC [95% CI] |
| --- | --- | --- | --- |
| Bórquez et al. (2023) | 0.97 [0.96 - 0.98] | 0.85 [0.79 - 0.90] | 0.98 [0.96 - 0.99] |
| Fan et al. (2024) | 0.97 [0.96 - 0.98] | 0.80 [0.71 - 0.87] | 0.98 [0.96 - 0.99] |
| Jung et al. (2024) | 0.97 [0.96 - 0.98] | 0.81 [0.72 - 0.88] | 0.98 [0.96 - 0.99] |
| Kabir et al. (2024) | 0.98 [0.97 - 0.99] | 0.80 [0.69 - 0.88] | 0.98 [0.96 - 0.99] |
| Mirimoghaddam et al. (2024) | 0.97 [0.96 - 0.98] | 0.80 [0.70 - 0.87] | 0.98 [0.96 - 0.99] |
| Mukundan (2019) | 0.98 [0.96 - 0.98] | 0.80 [0.71 - 0.87] | 0.98 [0.96 - 0.99] |
| Oliveira et al. (2020) | 0.98 [0.96 - 0.98] | 0.82 [0.73 - 0.88] | 0.98 [0.96 - 0.99] |
| Palm et al. (2023) | 0.98 [0.97 - 0.99] | 0.82 [0.72 - 0.89] | 0.98 [0.97 - 0.99] |
| Pedraza et al. (2024) | 0.98 [0.96 - 0.98] | 0.80 [0.70 - 0.87] | 0.98 [0.96 - 0.99] |
| Pham et al. (2023) | 0.97 [0.96 - 0.98] | 0.82 [0.73 - 0.88] | 0.98 [0.96 - 0.99] |
| Qaiser et al. (2018) | 0.97 [0.96 - 0.98] | 0.83 [0.75 - 0.89] | 0.98 [0.96 - 0.99] |
| Sode et al. (2023) | 0.98 [0.96 - 0.98] | 0.82 [0.73 - 0.88] | 0.98 [0.96 - 0.99] |
| Yao et al. (2022) | 0.97 [0.96 - 0.98] | 0.82 [0.73 - 0.88] | 0.98 [0.96 - 0.99] |
| **Combined** | **0.97 [0.96 - 0.98]** | **0.82 [0.73 - 0.88]** | **0.98 [0.96 - 0.99]** |

### 8. Adapted Risk of Bias and Applicability Concerns (QUADAS-AI)

**Figure S3:**
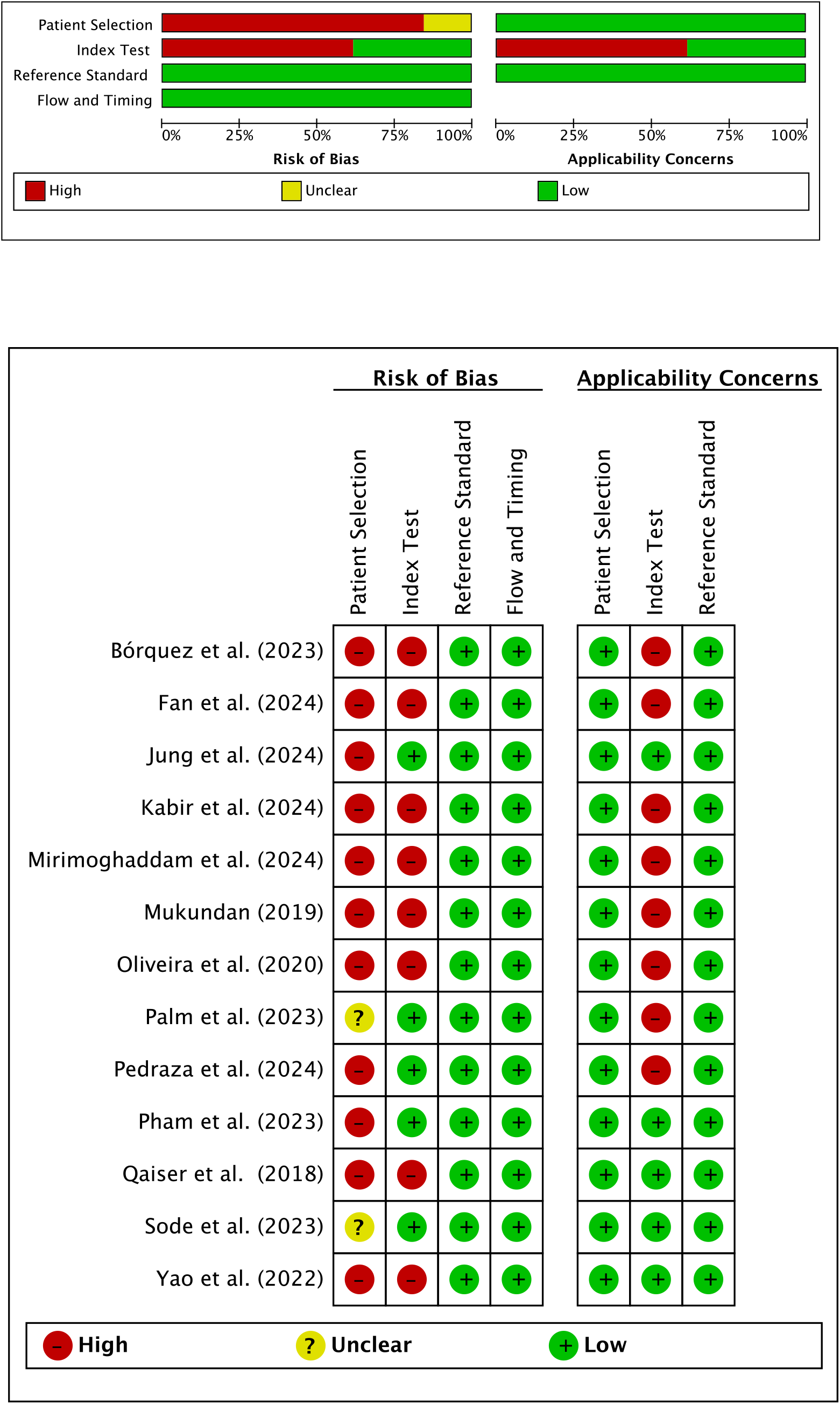
Risk of bias and applicability concerns of the included studies using the adapted QUADAS-AI tool.

## Footnotes

1 It is noteworthy that, for comparison purposes, we swapped the values of sensitivity and specificity, as the original criterion for positivity in their study was set to score 0, whereas in this review the positivity threshold was set to score 1+, 2+ or 3+.

## Notes

### Competing Interest Statement

The authors have declared no competing interest.

### Funding Statement

This study did not receive any funding.

### Author Declarations

The study used (or will use) ONLY openly available human data that were originally located at: MEDLINE (https://pubmed.ncbi.nlm.nih.gov), EMBASE (https://www.embase.com/landing?status=grey), Scopus (https://www.scopus.com/home.uri) and Web of Science (https://clarivate.com/academia-government/scientific-and-academic-research/research-discovery-and-referencing/web-of-science/) databases.

